# Transcatheter Aortic Valve Replacement Use in Young Patients Before and After Low-Risk Indication Approval

**DOI:** 10.1101/2024.05.22.24307638

**Authors:** Christina Waldron, Luigi Pirelli, Isaac George, Hiroo Takayama, Arnar Geirsson, Roland Assi, Makoto Mori

## Abstract

Transcatheter aortic valve replacement (TAVR) use in young patients has recently increased following FDA approval for use in low-risk patients. How TAVR use among young patients may have accelerated in the interval between the FDA approval and the guideline publication remain unknown. We sought to characterize the national trends in aortic valve replacement (AVR) among young patients before and after the low-risk indication approval. Using the National Inpatient Sample data, we conducted a cross-sectional study of patients younger than 65 years of age who underwent TAVR, SAVR, or Ross operations between January 1, 2016, and February 29, 2020. We compared in-hospital mortality before and after the low-risk indication approval to infer whether the expansion occurred among lower-risk strata within the TAVR group in relation to SAVR. We identified 106,340 AVRs, including 13,095 TAVR (12.3%), 63,620 bioprosthetic SAVR (59.8%), 28,370 mechanical SAVR (26.7%), and 1,255 Ross (1.2%). The mean age was 54 (10.9), including 32,775 (30.8%) women. Before and after the low-risk approval, the TAVR share increased at 0.25 (0.03%) and 0.60 (0.09%) per month, respectively (interaction term p-value <0.001). Expansion of TAVR use among young patients around the time of FDA approval in low surgical risk patients serves as a case study to highlight the potential importance of specifying the indicated age group in future FDA approvals of transcatheter valve intervention devices.

## Introduction

Transcatheter aortic valve replacement (TAVR) use in young patients has recently increased^1^. The U.S. Food and Drug Administration (FDA) approval of balloon-expanded and self-expandable TAVR devices for low-risk patients did not specify the indicated age group, although the participants in both pivotal trials, PARTNER 3 and Evolut low-risk trials, were old at a mean age of 73^2,1^. The high-risk nature of TAVR explant and unknown long-term valve durability are important considerations related to TAVR use among young patients with expected longevity^3^. While the current U.S. guideline recommends surgical aortic valve replacement (SAVR) in low-risk patients younger than 65^4^, the first guideline specifying such age threshold was published in 2020, 2 years after the first FDA approval of TAVR for low-risk patients. How TAVR use among young patients may have accelerated in the interval between the FDA approval and the guideline publication remain unknown.

Numerous trials are underway to evaluate transcatheter valve repair and replacement in non-aortic positions. With expected data lag and guideline publication lag accounting for the device durability and long-term outcomes, examining the rapidity of TAVR use expansion in young patients around the time of low-risk TAVR approval may serve as a case study to inform the nuances of future FDA approval of devices currently undergoing trials^5^.

To understand how the low-risk indication approval may have accelerated TAVR utilization nationally in this age group, we characterized the national trends in aortic valve replacement (AVR) among young patients before and after the low-risk indication approval.

## Methods

Using the National Inpatient Sample (NIS) data, a 20% stratified sample of all inpatient discharges from US non-federal hospitals, we conducted a cross-sectional study of patients younger than 65 years of age who underwent TAVR, SAVR, or Ross operations between January 1, 2016, and February 29, 2020 (the latest available year, up to the COVID-19 stay-at home order). Operations and aortic valve pathology were defined using the *International Classification of Diseases, 10*^*th*^ *revision* (ICD-10) (Supplemental Table). Patients with endocarditis were excluded. Difference-in-difference analysis estimated the association between the low-risk indication approval on August 16, 2018 (cutoff on September 1, 2018) and the change in the monthly percent TAVR share among all AVRs as a continuous variable (Supplemental Methods). We compared in-hospital mortality before and after the low-risk indication approval to infer whether the expansion occurred among lower-risk strata within the TAVR group in relation to SAVR.

We used the ‘survey’ R package, applied the hospital discharge weight variable in the NIS dataset to estimate the total national volume of index operations, and performed the chi-squared test, accounting for the sampled nature of the dataset. Difference-in-difference model was specified with the outcome being the monthly percent of TAVR share among all AVRs, with the covariates of the month as a nominal variable and a pre/post indicator of before and after September 1, 2018. An interaction term was added as the product of the month and pre/post indicator variable.

Analyses were performed in R version 4.2.2 (R Foundation). All P values were 2-sided, with <0.05 denoting statistical significance. This study was approved by the Yale Institutional Review Board. Informed consent was waived because the data were deidentified.

## Results

We identified 106,340 AVRs, including 13,095 TAVR (12.3%), 63,620 bioprosthetic SAVR (59.8%), 28,370 mechanical SAVR (26.7%), and 1,255 Ross (1.2%). The mean age was 54±10.9, including 32,775 (30.8%) women. The proportion of TAVR increased from 6.9% in 2016 to 22.4% in 2020. The shares of mechanical SAVR remained stable from 29.8% to 26%, while bioprosthetic SAVR decreased from 62.2% to 50.9%.

Before and after the low-risk approval, the TAVR share increased at 0.25±0.03% and 0.60±0.09% per month, respectively (interaction term *p*-value <0.001, Figure 1A). Among patients undergoing TAVR, in-hospital mortality was 2.4% before and 1.1% after the approval (*p*=0.015, Figure 1B). Among patients undergoing bioprosthetic SAVR, in-hospital mortality was 2.5% before and 2% after (*p*=0.1), and for mechanical SAVR, in-hospital mortality was 3% before and 3.5% after the approval (*p*=0.28). In-hospital mortality for patients undergoing the Ross procedure was 1.9% before and 1.1% after approval (*p*=0.67).

**Figure 1:**
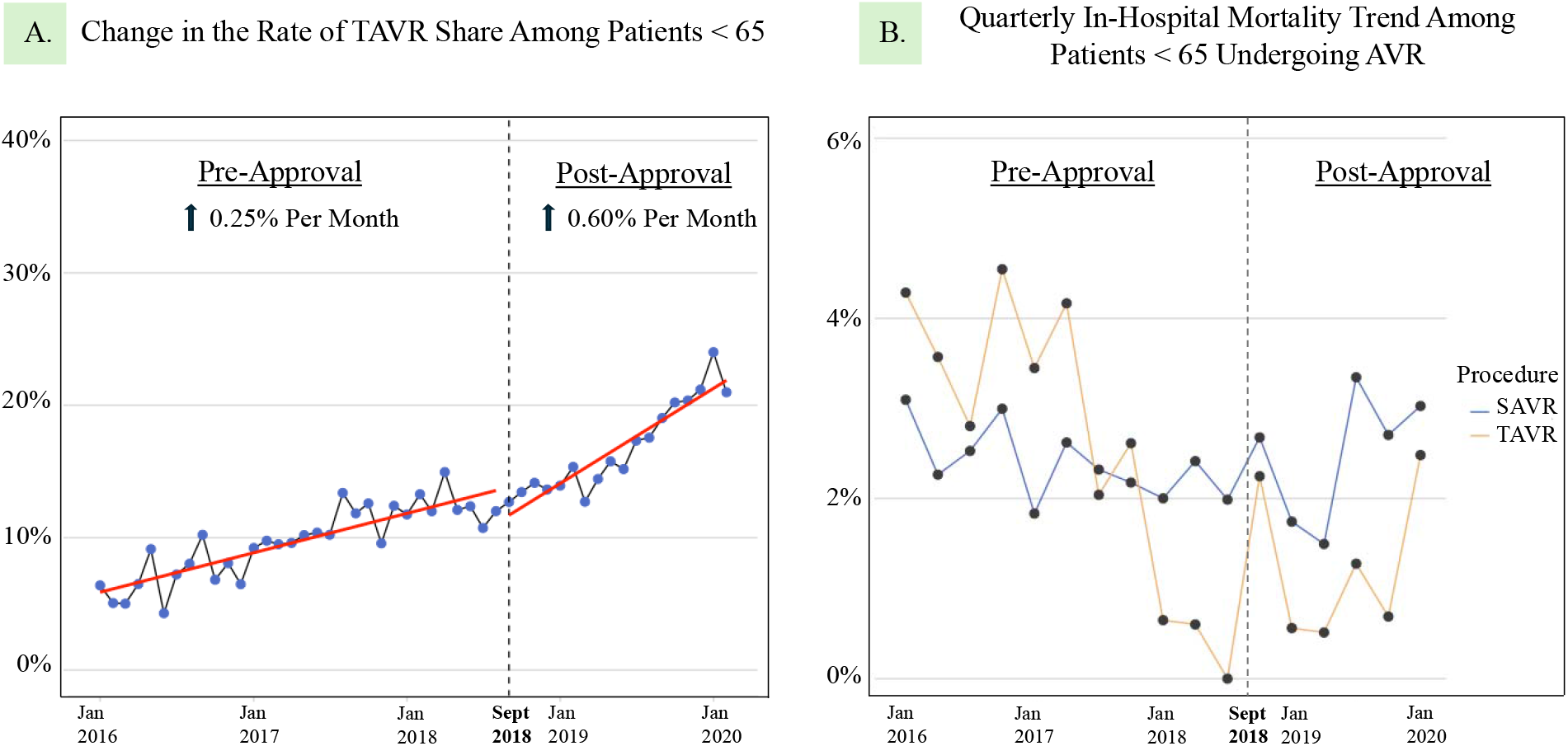
Annual Trends of Transcatheter Aortic Valve Replacement Shares and AVR Mortality Among Patients < 65 This figure shows a change in the rate of TAVR share among patients < 65 years of age before and after the first low-risk indication approval in August 2018 (panel A) and the quarterly in-hospital mortality trends among patients < 65 undergoing SAVR and TAVR (panel B). Red lines are linear regression lines fitted to the TAVR share before and after September 1, 2018. AVR: Aortic valve replacement; SAVR: Surgical aortic valve replacement; TAVR: Transcatheter aortic valve replacement.

Among patients younger than 50, TAVR share increased from 3.1% in 2016 to 6.7% in 2020. SAVR was increasingly performed for congenital pathology, including bicuspid valve (18% of SAVR in 2016 to 29% in 2020), while the pathology composition remained stable for TAVR.

## Discussion

We demonstrated that TAVR use in patients younger than 65 was increasing prior to the FDA approval for low-risk indication, and accelerated after the FDA approval, more than doubling the rate of monthly increase. The share of TAVR among all AVRs in this age group tripled between 2016 and 2020 preceding the publication of the first guideline specifying age threshold for low-risk TAVR. Significantly lower in-hospital mortality after the approval was observed only among TAVR patients, suggesting that the TAVR use in this young age group expanded towards lower-risk patients. These data suggest the importance of regulatory bodies to delineate the indicated age group for novel indication transcatheter valve devices, as the first guidance specifying the age threshold lagged by 2 years following this FDA approval.

Considerations surrounding the patient age is relevant, because low risk is not synonymous to young age in prosthetics with age-dependent durability. Yet, the accelerated increase in TAVR share among young patients around the time of the FDA approval may reflect the public’s reception of the low-risk approval. Ongoing trials on moderate or asymptomatic severe aortic stenosis may follow similar trend^6^. Similarly, transcatheter edge-to-edge mitral repair is being compared with surgical mitral valve repair^7^. The trial population will likely be old, with the inclusion criteria of older than 65, which is above the national mean of those undergoing surgical MV repair for degenerative disease^7^. With less competing risks, the long-term impact of residual mitral regurgitation and the impact of device durability are likely more pronounced in younger patients. As novel devices expand in indications, the momentum of ‘indication creep’^5^ may be balanced with a clear age threshold for indication by the regulatory bodies.

The NIS is claims-based and lacks granular risk characteristics. Therefore, we used in-hospital mortality as a surrogate of patient risk profile. Valve pathology diagnosis was missing in 15%.

## Conclusion

Expansion of TAVR use among young patients around the time of the FDA approval of balloon-expandable TAVR in low surgical risk patients serves as a case study to highlight the potential importance of specifying the indicated age group in future FDA approvals of transcatheter valve intervention devices that may have age-dependent durability and unknown long-term implications.

## Data Availability

The data will be available upon reasonable request to the corresponding author.

---

**Supplementary Table:** ICD Codes for Procedures and Aortic Pathology Etiologies

| Procedure or Aortic Pathology | ICD Code |
| --- | --- |
| TAVR | 02RF37Z, 02RF38Z, 02RF3JZ, 02RF3KZ |
| Bioprosthetic/biologic SAVR | 02RF07Z, 02RF08N, 02RF08Z, 02RF0KZ,<br>02RF48N, 02RF48Z, 02RF4KZ, 02RF47Z |
| Mechanical SAVR | 02RF0JZ, 02RF4JZ |
| Ross Operation<sup>1</sup> | Defined as simultaneous use of biological<br>AVR and replacement of pulmonary valve<br>with tissue graft, including homograft |
| Biologic AVR | 02RF07Z, 02RF08Z, 02RF0KZ |
| Replacement of pulmonary Valve with<br>tissue graft, including homograft | 02RH07Z, 02RH08Z, 02RH0KZ, 02RH47Z |
| Infective Endocarditis | I33, I38, I339, T826, B376, I39, I398, I423,<br>I091, I011, A3951, A3282, B3321, A5483 |
| Aortic stenosis | I350 |
| Aortic insufficiency | I351 |
| Mixed aortic stenosis and insufficiency | I352 |
| Congenital AI/AS including BAV | Q2383, Q231, Q230 |

AI: Aortic Insufficiency; AS: Aortic Stenosis; BAV: Bicuspid Aortic Valve; SAVR: Surgical
Aortic Valve Replacement; TAVR: Transcatheter Aortic Valve Replacement.

## References

1. Sharma T, Krishnan AM, Lahoud R, Polomsky M, Dauerman HL. National Trends in TAVR and SAVR for Patients With Severe Isolated Aortic Stenosis. Journal of the American College of Cardiology. 2022;80(21):2054–2056. doi:10.1016/j.jacc.2022.08.787

2. Mack MJ, Leon MB, Thourani VH, et al. Transcatheter Aortic-Valve Replacement with a Balloon-Expandable Valve in Low-Risk Patients. New England Journal of Medicine. 2019;380(18):1695–1705. doi:10.1056/NEJMoa1814052

3. Hawkins RB, Deeb GM, Sukul D, et al. Redo Surgical Aortic Valve Replacement After Prior Transcatheter Versus Surgical Aortic Valve Replacement. JACC: Cardiovascular Interventions. 2023/04/24/ 2023;16(8):942–953. doi:10.1016/j.jcin.2023.03.015

4. Otto CM, Nishimura RA, Bonow RO, et al. 2020 ACC/AHA Guideline for the Management of Patients With Valvular Heart Disease: A Report of the American College of Cardiology/American Heart Association Joint Committee on Clinical Practice Guidelines. Circulation. 2021;143(5):e72–e227. doi:10.1161/CIR.0000000000000923

5. Goel SS, Reardon MJ. Indication Creep in Transcatheter Aortic Valve Implantation—Data or Desire? JAMA Cardiology. 2023;8(6):519–520. doi:10.1001/jamacardio.2023.0674

6. PROGRESS: Management of Moderate Aortic Stenosis by Clinical Surveillance or TAVR (PROGRESS). ClinicalTrials.gov Identifier: NCT04889872. Updated April 29, 2024. Accessed May 19, 2024. https://classic.clinicaltrials.gov/ct2/show/NCT04889872.

7. McCarthy PM, Whisenant B, Asgar AW, et al. Percutaneous MitraClip Device or Surgical Mitral Valve Repair in Patients With Primary Mitral Regurgitation Who Are Candidates for Surgery: Design and Rationale of the REPAIR MR Trial. Journal of the American Heart Association. 2023;12(4):e027504. doi:10.1161/JAHA.122.027504

## References

1. El-Hamamsy I, Toyoda N, Itagaki S, et al. Propensity-Matched Comparison of the Ross Procedure and Prosthetic Aortic Valve Replacement in Adults. J Am Coll Cardiol. 2022;79:805–815.

